# Development of a clinical model to predict vagus nerve stimulation response in pediatric patients with drug-resistant epilepsy

**DOI:** 10.1101/2022.01.13.22269237

**Authors:** Nallammai Muthiah, Arka N. Mallela, Lena Vodovotz, Nikhil Sharma, Emefa Akwayena, William Welch, George M. Ibrahim, Taylor J. Abel

## Abstract

**Introduction:** Epilepsy impacts 470,000 children in the United States, and children with epilepsy are estimated to expend 6 times more on healthcare than those without epilepsy. For patients with antiseizure medication (ASM)-resistant epilepsy and unresectable seizure foci, vagus nerve stimulation (VNS) is a treatment option. Predicting response to VNS has been historically challenging. We aimed to create a clinical prediction score which could be utilized in a routine outpatient clinical setting.

**Methods:** We performed an 11-year, single-center retrospective analysis of patients <21 years old with ASM-resistant epilepsy who underwent VNS. The primary outcome was ≥50% seizure frequency reduction after one year. Univariate and multivariate logistic regressions were performed to assess clinical factors associated with VNS response; 70% and 30% of the sample were used to train and validate the multivariate model, respectively. A prediction score was developed based on the multivariate regression. Sensitivity, specificity, and area under the receiver operating curve (AUC) were calculated.

**Results:** This analysis included 365 patients. Multivariate logistic regression revealed that variables associated with VNS response were: <4 years of epilepsy duration before VNS (p=0.008) and focal motor seizures (p=0.037). The variables included in the clinical prediction score were: epilepsy duration before VNS, age at seizure onset, number of pre-VNS ASMs, if VNS was the patient’s first therapeutic epilepsy surgery, and predominant seizure semiology. The final AUC was 0.7013 for the “fitted” sample and 0.6159 for the “validation” sample.

**Conclusions:** We developed a clinical model to predict VNS response in one of the largest samples of pediatric VNS patients to date. While the presented clinical prediction model demonstrated an acceptable AUC in the training cohort, clinical variables alone likely do not accurately predict VNS response. This score may be useful upon further validation, though its predictive ability underscores the need for more robust biomarkers of treatment response.

## Introduction

In the United States, approximately 470,000 children have active epilepsy.^1^ Of these children, it is estimated that 30% will have seizures resistant to antiseizure medications (ASMs).^2^ For patients with multifocal or generalized epilepsy, focal epilepsy in an eloquent brain region, or when intracranial procedures are infeasible secondary to medical contraindications, neurosurgical seizure focus resection may not be an option. Vagus nerve stimulation (VNS) is a potential treatment for these patients. Approximately 1,000-2,000 VNS procedures are performed each year^3^, and today, about 30,000 pediatric patients have a vagus nerve stimulator for seizure frequency control.^4^

VNS is a common treatment for drug-resistant epilepsy. Randomized controlled trials in adults demonstrated that VNS is safe and effective at reducing seizure frequency, especially at higher stimulation parameters.^5-8^ In these initial studies, the primary outcome used to assess VNS effectiveness was >50% seizure frequency reduction at ≥3 months compared to baseline.^5^ Since then, this outcome has been utilized widely in VNS literature. Today, it is estimated that between 45% and 65% of patients achieve at least 50% seizure frequency reduction with VNS.^9-11^ In the pediatric population, the effectiveness of VNS is more varied, with reported rates between 20% and 70%.^12-15^

Predicting VNS response remains elusive. Workewych et al. have suggested that brain connectivity is a promising avenue to predict outcomes following VNS.^16^ However, these techniques are under development and most epilepsy centers do not have network-based connectomic biomarkers readily available during initial discussions of VNS therapy. We sought to evaluate whether a simple prediction model based on clinical factors alone could guide initial discussions about the individualized benefits of VNS therapy for a given patient with ASM-resistant epilepsy.

We performed an 11-year retrospective analysis of pediatric patients with drug-resistant epilepsy who underwent VNS at UPMC Children’s Hospital of Pittsburgh. The objectives of the present study were twofold. First, we sought to assess the effectiveness of VNS in our pediatric patient sample. Second, we aimed to develop a clinical prediction tool to predict which patients with ASM-resistant epilepsy would respond to VNS after one year of therapy.

## Methods

### Data acquisition

We conducted a retrospective analysis of all VNS implantations at UPMC Children’s Hospital of Pittsburgh from 2009 to 2020. Patient demographics were collected using the electronic medical record. Clinical neurology and neurosurgery notes were reviewed to determine pre-VNS epilepsy-related variables including: seizure semiology^17^, seizure etiology (classified as “structural”, “genetic”, “metabolic”, “infectious”, “immune”, and “unknown”^18^), age at seizure onset, seizure frequency before VNS (categorized as “less than one seizure per month”, “at least one seizure per month but less than one seizure per week”, “at least one seizure per week but less than one seizure per day”, “at least one seizure per day but less than one seizure per hour”, and “at least one seizure per hour”), and number of pre-VNS ASMs. Pre-VNS data reflected the most recent clinical note available prior to VNS implantation and did not exceed 12 months before VNS implantation. Change in overall seizure frequency from baseline (coded as “<50% seizure frequency reduction”, “50% seizure frequency reduction”, and “>50% seizure frequency reduction”) were collected using clinical neurology and neurosurgery notes at approximately one year after VNS implantation. There was some variability in follow-up appointment scheduling, so data were collected as close to the one-year time point as possible.

The primary outcome for this analysis was response to VNS therapy (measured as ≥50% reduction in average predominant seizure frequency) at one year. If patients had seizures or seizure clusters clearly associated with an exogenous trigger—specifically any recent or active infection, clear lack of sleep, uncharacteristic dehydration, or uncharacteristic medication non-adherence in the days-to-week before the seizure/seizure cluster—these seizures were not counted towards the patients’ average seizure frequency at one year.

### Study participants

Patients were included in demographic and seizure characterization analysis if they had ASM-resistant epilepsy and underwent VNS implantation at any age <21 years. ASM-resistant epilepsy was defined as “the failure of adequate trials of two tolerated, appropriately chosen and used antiepileptic drug schedules to achieve sustained seizure freedom.”^19^ Patients were excluded from primary outcome analysis if their VNS was removed or permanently turned off within 12 months of initial implantation (n=4). Patients who had their VNS permanently turned off or removed were still included in demographics analysis.

### Statistical analyses

The primary outcome for this analysis, VNS response, was coded as a binomial variable. Patients were considered to have VNS response if they achieved at least 50% seizure frequency reduction in their predominant seizure semiology at one year after VNS implantation. The sample was randomly divided into a “training set” and “validation” set. The training set consisted of 70% of the sample, while the validation set was comprised of the remaining 30%. The training set was used for multivariate logistic regression model development and the validation set was used to test the fit, discrimination, and calibration of the resulting model. Chi-squared and independent sample t-tests were performed to assess baseline demographic and clinical differences between the training and validation samples. *Supplementary Figure 1* demonstrates the patient inclusion and exclusion criteria as well as the sample splitting procedure.

Univariate logistic regressions were performed to assess the association between each of the above predictors with VNS response at one year. The variables which were considered for inclusion in the multivariate model were those 1) selected by an initial LASSO regression for model regularization or 2) believed to be clinically important indicators of patients’ response to VNS for drug-resistant epilepsy based on evidence provided by literature: age at epilepsy onset, duration of epilepsy before VNS implantation, predominant seizure semiology before VNS, etiology of epilepsy, number of pre-VNS ASMs, and prior therapeutic epilepsy surgeries. The variables included in the final multivariate model were selected by assessing receiver operative curve characteristics. Multicollinearity was assessed with variance inflation factors, with values greater than 10 considered to be evidence of multicollinearity.

After fitting the final model, data were subsequently screened for influential points and outliers. Influential points were determined by calculating Pregibon’s Δß for each patient, with absolute values greater than 2 considered to be influential. Similarly, outliers were determined by calculating standardized Pearson residuals, with absolute values greater than 2 considered to be outliers. Outliers and influential points were compared to non-outliers and non-influential points, respectively, with chi-2 tests to assess whether VNS response significantly differed; p<0.05 was considered significant.

### Model validation

The adequacy of the final model was tested in multiple ways. First, the predicted probability of VNS response at one year between those who did and did not achieve VNS response at one year was compared using an independent sample t-test. Multivariate model fit was also assessed with the Hosmer-Lemeshow goodness of fit test, with a p-value<0.05 considered to be indicative of poor model fit. Model discrimination was assessed by determining the area under the receiver operating characteristic curve (AUC), a measure of concordance which is equivalent to the C-statistic for binary outcomes.^20^ The model was validated by recalculating these statistics using the validation sample, given the parameters estimated from the training sample. The model was also validated with crossfold validation in which the sample was randomly re-split (80% training, 20% validation) into five independent samples.^21^

### Clinical prediction score calculation

Clinical score development followed the steps outlined by Sullivan et al.^22^ The parameters of the model were estimated with the final multivariate logistic regression, as mentioned above. Risk factors were organized into categories. All non-binomial categorical variables were categorized into ordinal or categorical variables based on the spread of our sample data. Epilepsy duration before VNS was categorized into: 0-3 years, 4-8 years, 9-12 years, and >12 years. Predominant seizure semiology was categorized based on the 2017 ILAE recommendations as focal motor, focal non-motor, focal to tonic clonic, generalized motor, or generalized non-motor seizures. Very few patients in our cohort had “unknown” predominant seizure origins, so these patients were excluded from mutivariate model fitting. Number of pre-VNS ASMs was categorized into 0-1, 2-3, or 4-5 ASMs. Reference categories were selected such that other coefficients for a given variable were positive (i.e., all non-reference categories were associated with higher odds of VNS response than the reference). Whether a patient underwent pre-VNS therapeutic epilepsy surgery was coded as a binary variable. The referent risk factor profile was determined selecting the base category for each risk factor and assigning the “risk” score 0 to the reference category. The distance in regression units for each non-reference category within a given risk factor was calculated. Each regression coefficient was converted to an integral score using a fixed multiplier. Finally, the risk associated with each possible combination of point totals was determined using the logistic function.^22^

Outcomes of the prognostic multivariate logistic regression model are reported in concordance with the Transparent Reporting of a multivariable prediction model for Individual Prognosis Or Diagnosis (TRIPOD) guidelines.^23^

## Results

This analysis included 365 patients of whom 339 had outcome data at one year. Total sample demographics are listed in left-most column of Table 1. The remainder of Table 1 displays the demographic and pre-VNS epilepsy characteristics among the training and validation samples. There were no clinically or statistically significant differences between training and validation samples. As expected, patient demographics were split as approximately 70% and 30% between the training and validation samples, respectively. Overall, 48% of patients (n=163) responded to VNS at one year. Univariate logistic regressions (see *Supplementary Table 1*) revealed that variables associated with VNS response were: epilepsy duration before VNS of 0-3 years (OR=2.90, 95% CI=[1.2-7.1], p=0.019) and age ≥3 years at epilepsy onset (OR=1.63, 95% CI=[1.1-2.5], p=0.028).

**Table 1.** Demographics and pre-vagus nerve stimulator (VNS) epilepsy characteristics of the total study sample, the training sample (a randomly selected 70% of the total sample), and the validation sample (the remaining randomly selected 30% of the total sample). VNS: vagus nerve stimulation, ASMs: antiseizure medications. p<0.05 was considered significant. Categorical variables reported as n(%), continuous variables reported as mean ± standard deviation, and number of ASMs reported as mean [25^th^ percentile, 75^th^ percentile]. ^F^Fisher’s exact test ^W^Wilcoxon rank-sum test All tests were either χ^2^ tests or independent sample t-tests assuming equal variances unless otherwise noted. Non-parametric tests were used when appropriate; p<0.05 was considered significant

|  | Total sample<br>(n=365) | Training sample<br>(n=255) | Validation sample<br>(n=110) | p-value |
| --- | --- | --- | --- | --- |
| Sex (F) | 177 (48.5%) | 123 (49.1%) | 54 (50.9%) | 0.881 |
| Etiology of epilepsy |  |  |  | 0.902<sup>F</sup> |
| Structural | 120 (33.2%) | 81 (32.1%) | 39 (35.8%) |  |
| Genetic | 95 (26.3%) | 67 (26.6%) | 28 (25.7%) |  |
| Metabolic | 3 (0.8%) | 2 (0.8%) | 1 (0.9%) |  |
| Infectious | 11 (3.0%) | 9 (3.6%) | 2 (1.8%) |  |
| Unknown | 132 (36.6%) | 93 (36.9%) | 39 (35.8%) |  |
| Seizure semiology |  |  |  | 0.807 |
| Focal motor | 86 (26.5%) | 58 (26.0%) | 28 (27.5%) |  |
| Focal non-motor | 51 (15.7%) | 28 (17.0%) | 13 (12.8%) |  |
| Focal to tonic clonic | 41 (12.6%) | 27 (12.1%) | 14 (13.7%) |  |
| Generalized motor | 110 (33.9%) | 73 (32.7%) | 37 (36.3%) |  |
| Generalized non-motor | 37 (11.4%) | 27 (12.1%) | 10 (9.8%) |  |
| Epilepsy type |  |  |  | 0.506 |
| Focal | 170 (48.7%) | 121 (50.2%) | 49 (45.3%) |  |
| Generalized | 114 (32.7%) | 74 (30.7%) | 40 (37.0%) |  |
| Combined focal/generalized | 65 (18.6%) | 46 (19.1%) | 19 (17.6%) |  |
| Patient deceased | 16 (4.1%) | 13 (5.1%) | 3 (2.8%) | 0.411<sup>F</sup> |
| Age at seizure onset (years) | 3.3 ± 4.0 | 3.0 ± 4.0 | 3.8 ± 4.2 | 0.113<sup>w</sup> |
| Epilepsy duration pre-VNS (years) | 6.5 ± 4.0 | 6.8 ± 4.2 | 5.9 ± 3.4 | 0.164<sup>w</sup> |
| Quantity of pre-VNS ASMs | 2.2 [2, 3] | 2.2 [2, 3] | 2.3 [2, 3] | 0.940<sup>w</sup> |
| VNS response at one year | 163 (48.1%) | 112 (47.5%) | 51 (49.5%) | 0.727 |

Within the training sample (n=255), the final multivariate logistic regression included: epilepsy duration before VNS, age at seizure onset, number of pre-VNS ASMs, if VNS was the patient’s first therapeutic epilepsy surgery, and predominant semiology (Table 2). The Hosmer-Lemeshow test was not significant (p=0.112), indicating no statistical evidence for lack of model fit. All variance inflations factors were <4, with an average variance inflation factor of 2.39. Therefore, we determined that there was no evidence to suggest multicollinearity among variables in the model.

**Table 2.**
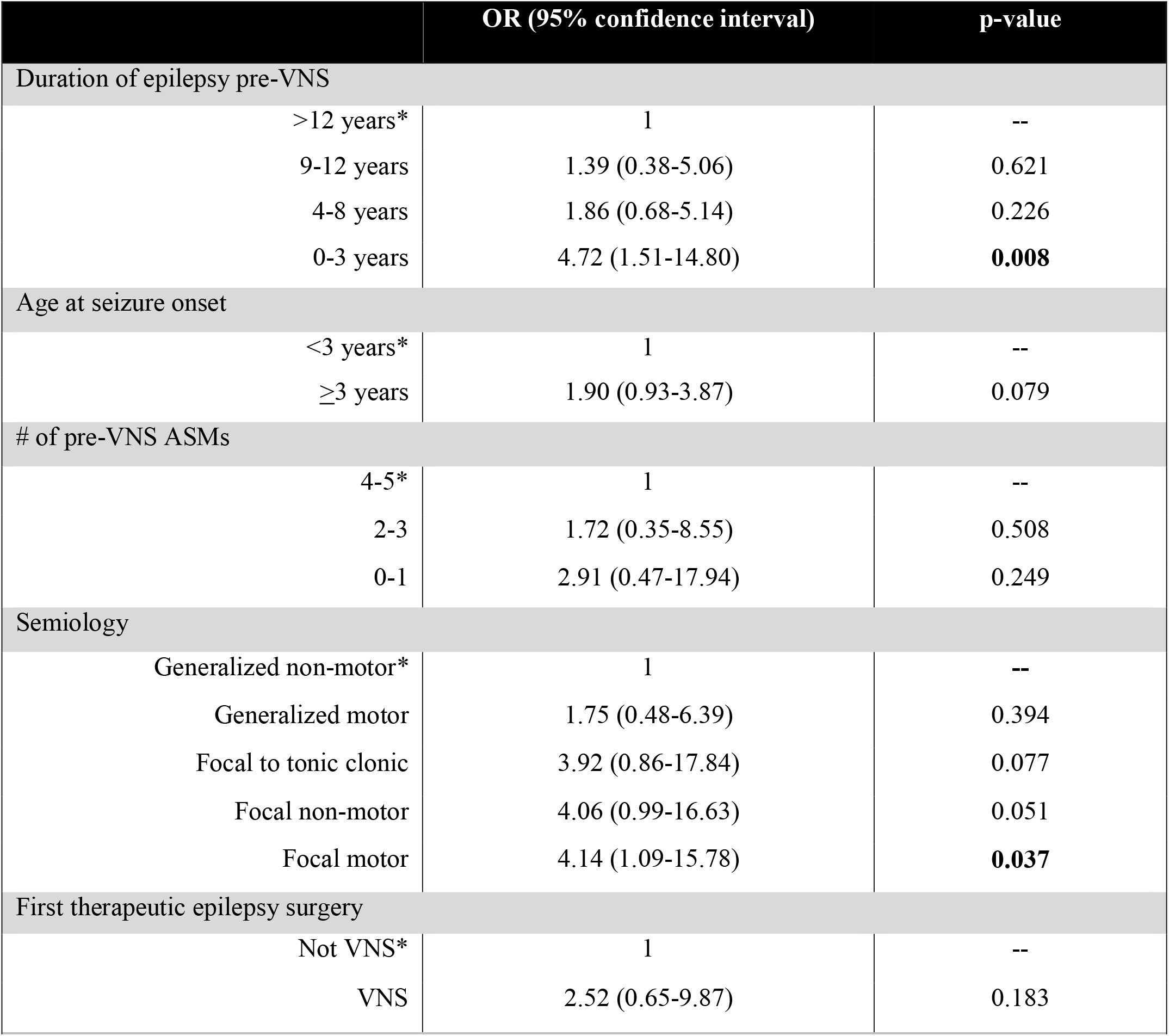
Odds ratios (ORs) and p-values from the final multivariate logistic regression evaluating the association between respective covariates and VNS response at one year, fitted on the total sample. Higher ORs are indicative of higher odds of VNS response. VNS: vagus nerve stimulation, ASMs: antiseizure medications. p<0.05 was considered significant. *designates reference category

Within the training sample, the model demonstrated an AUC of 0.70 (Figure 1). In the validation sample, the model demonstrated an AUC of 0.62 (Figure 1). After performing five-fold, “leave-one-out” validation, the mean validation AUC was 0.65 (range: 0.55 to 0.71), with a 95% bootstrap bias corrected confidence interval of 0.6019, 0.7531. Given that an AUC of ≥0.70 is considered acceptable for model discrimination, our AUC was acceptable in the training sample but not in the validation sample. With a cutoff of 0.5, the model demonstrated a sensitivity of 65%, specificity of 64%, and correctly predicted VNS response in 65% of our sample.

**Figure 1.**
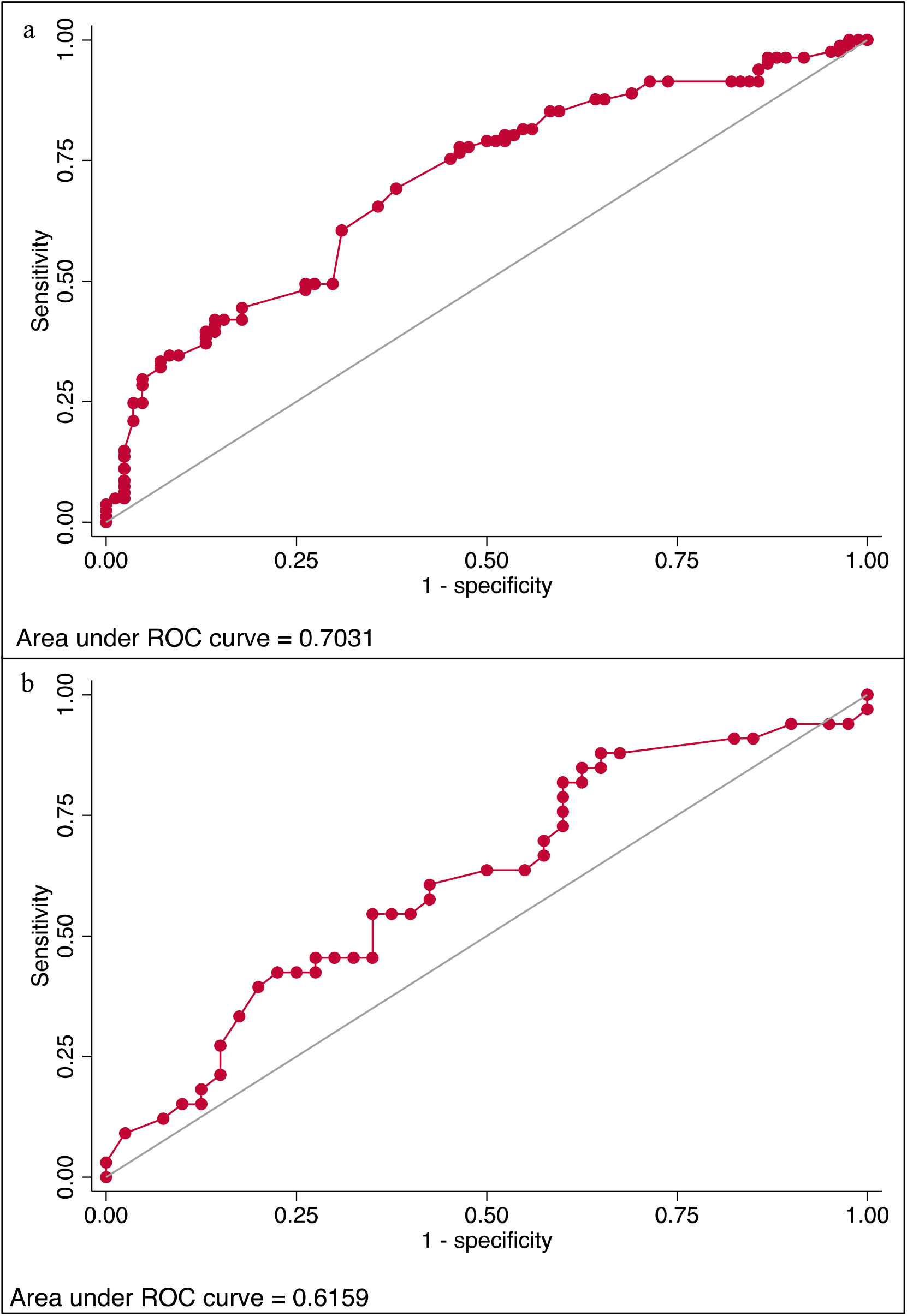
a) Training and b) validation cohort receiver operating characteristic (ROC) curves with respective areas under the ROC displayed in the bottom left.

The clinical prediction score was developed from this multivariate logistic regression model. The final score was out of 5 total points. Table 3 displays the scoring system for this clinical prediction tool. *Supplementary Table 2* delineates the number of patients in our sample who scored each point total, the probability of VNS response for each point total as estimated by the clinical tool, and the actual proportion of patients who achieved VNS response by one year. Most patients in this sample scored 2, 3, or 4 on the clinical score, while no patients scored a 0. The probabilities predicted by the VNS response score roughly approximated the actual VNS response (Figure 2), with a correlation coefficient of 0.81. The Hosmer-Lemeshow model calibration test demonstrated good logistic regression model calibration (*see Supplementary Figure 2*).

**Table 3.** VNS clinical prediction scoring system. Higher points represent a greater likelihood of VNS response by one year. The gray cells represent the base scenario, representing the characteristics associated with lowest probability of VNS response by one year. *Any other semiology includes generalized motor, focal non-motor, focal motor, and focal to tonic clonic

| Points | Epilepsy duration | Age at epilepsy onset | First therapeutic epilepsy surgery was VNS | # of ASMs | Semiology |
| --- | --- | --- | --- | --- | --- |
| 0 | $\geq 9$ years | $< 3$ years | No | 2-5 | Generalized Non-Motor |
| 1 | $< 9$ years | $\geq 3$ years | Yes | 0-1 | Any other semiology* |

**Figure 2.**
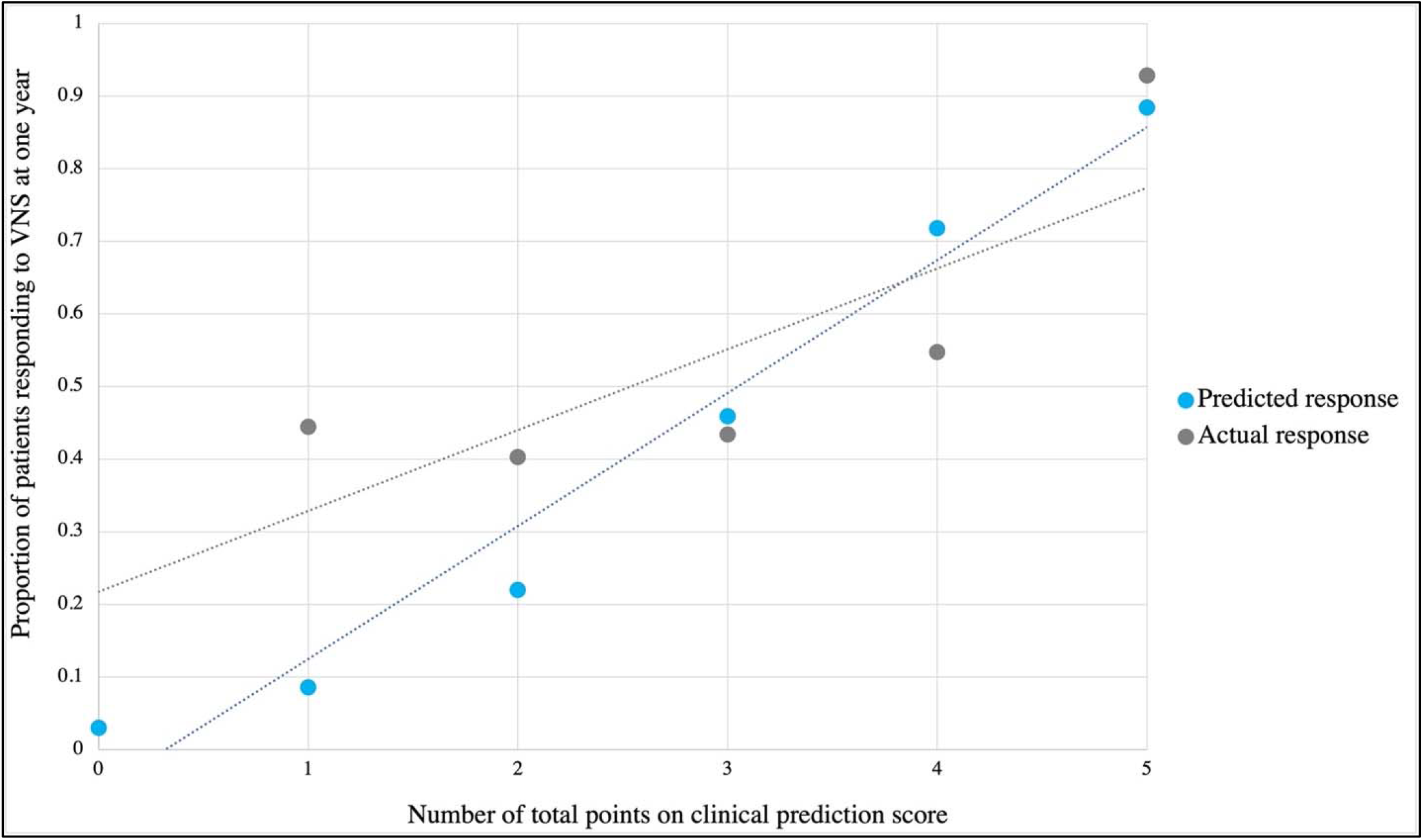
Actual proportion of patients responding to VNS by one year by point total compared to the predicted probabilities of VNS response by the clinical score.

## Discussion

In this analysis, we developed a clinical prediction model using a large single-center sample of pediatric VNS patients. This study’s proposed scoring instrument was based on a multivariate logistic regression model. The results of our model indicated that patients with epilepsy duration <4 years and patients with focal motor seizures were significantly more likely to respond to VNS at one year.

Our goal was to develop a prediction model using measurable clinical variables that would facilitate use in the outpatient setting. Though our model demonstrated an ability to predict VNS response at a rate slightly better than chance, our analysis demonstrated that clinical variables alone are not sufficient to accurately predict VNS response in this patient population. More advanced data, like biomarkers^16^, are warranted for more accurate predictions of VNS response. This tool may be utilized as an adjunct during the initial outpatient evaluation of seizure control options for pediatric patients with ASM-resistant epilepsy.

### Factors associated with VNS response at one year

Many studies have demonstrated the association between shorter duration of epilepsy and VNS response.^24-27^ A recent meta-analysis by Wang et al. demonstrated that among 504 patients from seven independent studies, those who responded to VNS exhibited a standardized mean difference of -0.22 years epilepsy duration compared to those who did not respond to VNS.^25^ Similarly, Zhu et al., in their retrospective review, reported that adults with a pre-surgery epilepsy duration of 2-5 years had approximately seven times the odds of response to VNS compared to those with an epilepsy duration of ≥12.5 years.^26^ Our results similarly demonstrated that patients with <4 years of epilepsy duration pre-VNS had 4.7 times the odds of responding to VNS by one year of stimulation than those with >12 years of epilepsy duration.

Our study also revealed that patients with predominantly focal motor seizures were significantly more likely to respond to VNS than those with predominantly generalized non-motor seizures. We observed a graded response: patients with focal seizures and patients with motor seizures had higher odds of achieving VNS response than those with generalized seizures and those with non-motor seizures, respectively. Taken in context of prior literature, these findings are not surprising. Englot et al. demonstrated that patients with predominantly focal seizures experienced 1.37 times the odds of VNS response at one year, while those with predominantly generalized tonic clonic seizures (either primarily or secondarily) exhibited 0.77 times the odds of VNS response at one year.^27^ Still, in their sample, 46-49% of patients with generalized tonic clonic seizures responded to VNS at one year,^27^ suggesting that patients with generalized seizures can still benefit from VNS. Given that VNS is currently only FDA-approved for patients with focal seizures, our results add to the existing pool of literature that VNS could be beneficial for patients with predominantly focal or generalized seizures.

We found that no etiologies of epilepsy were significantly associated with VNS response (presented in *Supplementary Table 1*). Evidence is mixed regarding epilepsy etiology and VNS response. Englot et al., in a meta-analysis, suggested that patients with tuberous sclerosis-related epilepsy were significantly more likely to respond to VNS than those with idiopathic or unknown causes.^27^ Other evidence also suggests that lesional epilepsy, post-traumatic epilepsy^27,28^, or post-stroke epilepsy^28^ could be associated with positive VNS response. In our analysis, only 24 and 12 patients had epilepsy secondary to stroke and trauma, respectively. Neither variable was significantly associated with VNS response (data not presented). These discrepant results may be because etiologies like “post-traumatic epilepsy”, “post-stroke epilepsy”, and “tuberous sclerosis complex” are heterogeneous. Perhaps the implicated brain circuitry is more associated with VNS response than the etiology itself.

In our analysis, predominant seizure phenotype (i.e., atonic, tonic, absence, etc.) and epilepsy type (focal, generalized, combined focal/generalized) were considered for inclusion in the final model. Both variables were ultimately omitted given that they demonstrated signs of overfitting and were likely to be multicollinear with seizure semiology. Still, these variables offer interesting avenues for further exploration. In a recent retrospective analysis of 41 patients by Boluk et al., patients with combined focal/generalized epilepsy experienced notably more seizures per day (median: 6/day) when compared to those with generalized epilepsy (median: 0.6.day) and focal epilepsy (median: 1.5/day).^29^ While all three groups exhibited significant decrease in seizure frequency after one year of VNS, those with combined focal/generalized epilepsy exhibited a 50% median seizure frequency decrease, while those with focal epilepsy exhibited a median seizure frequency decrease of 75%. Considering patients with combined focal/generalized epilepsy have multiple semiologies by nature, it is possible that certain semiologies tend to be resistant to VNS therapy^30,31^ and predominant seizure frequency went unchanged. For example, 20% of our patients with combined focal/generalized epilepsy had primarily atonic seizures pre-VNS (versus only 11% and 4% of generalized and focal epilepsy patients, respectively). Evidence suggests that atonic seizures are not optimally treated with VNS therapy.^30,32^ In their landmark meta-analysis, Lancman et al. demonstrated that only 26% of patients achieved >75% atonic seizure frequency reduction with VNS, whereas 70% of patients achieved >75% atonic seizure frequency reduction with corpus callosotomy.^30^

### Clinical predictive model

Predicting which patients will respond to VNS response has traditionally been challenging. Workewych et al. performed a systematic review of preoperative biomarkers which could predict response to VNS after one year of therapy. The authors demonstrated that heart rate variability was an independent marker of VNS response and their results indicated that brain network connectivity analyses are a promising avenue for predicting seizure responsiveness to VNS.^16^ Such connectonomic and biomarker analyses have an important role in understanding the mechanism of VNS and could aid patient selection at specialized epilepsy centers. The variables utilized in this study’s clinical prediction tool are easily attainable by an outpatient care provider and demonstrates better predictive accuracy (65%) than chance alone.

From our data, a score of 4 or more is associated with at least a 72% probability of VNS response, while a score of 3 or less is associated with a <46% probability of response. Interestingly, most patients in our sample scored a 2 (n=67), 3 (n=136), or 4 (n=84) out of 5 total points. Only 14 patients scored a 5. In other words, based on our score, a substantial proportion of patients in our sample were predicted to have at most, a 22% probability of VNS response by one year. In fact, among patients with a clinical score of 2, 40% responded to VNS at one year. This is consistent with the performance properties of our model (64% specificity and 65% sensitivity) and suggests that our instrument, as is, may be utilized primarily to exclude patients for VNS surgery. It is feasible that a version of this tool could be utilized in conjunction with appropriate seizure mapping and multidisciplinary discussion to selectively recommend VNS to pediatric patients.

Considering the broader context of modern epilepsy surgery, VNS is one of many options for patients with treatment-resistant epilepsy. Other treatment modalities include responsive neurostimulation (RNS) and deep brain stimulation (DBS). RNS, a closed loop neurostimulation system designed to abort seizure activity from the seizure onset zone,^33^ has been demonstrated to be effective for both pediatric and adult patients. While RNS is currently only approved by the Food and Drug Association (FDA) for adult patients with ASM-resistant epilepsy, small-scale studies have indicated that RNS is associated with both periods of seizure freedom as well as long-term, ≥80% seizure frequency reduction in pediatric patients.^34^ DBS is another alternative surgical treatment for ASM-resistant seizures. Evidence suggests that between 60% and 95% of patients can achieve >50% seizure frequency reduction with DBS, and the response rate appears to vary by location of electrode implantation (anterior nucleus of the thalamus vs. centromedian nucleus vs. subthalamic nucleus).^33,35^

### Limitations/Future work

This study has key limitations that must be carefully considered. Firstly, our study design is retrospective in nature. As such, it is subject to the selection and information biases which limit all retrospective studies. Secondly, our outcome data is based on parent report, which is subject to recall bias. Parents may have reported some clinical events that may not have been “true” seizures electrographically. Similarly, some “true” seizures may have gone unnoticed or were forgotten. We addressed this limitation in two ways. First, we categorized our outcome as a binary variable: <50% or ≥50% seizure frequency reduction from baseline. This allowed us to account for variations in the exact frequency of seizures. Second, we used the primary treating neurologist’s expertise whenever possible to quantify seizure frequency. Therefore, our data likely accurately reflected patients’ overall response to VNS. One final limitation is that this data is all based on one single epilepsy referral center. Balancing this limitation, our cohort is the largest pediatric VNS database to date. Still, future, multi-center studies would be an excellent avenue to increase sample size and improve the generalizability of our findings.

## Conclusions

We developed a clinical model to predict VNS response in one of the largest samples of pediatric VNS patients to date. While the presented clinical prediction model demonstrated an acceptable AUC in the training cohort, clinical variables alone likely do not accurately predict VNS response. This score may be useful upon further validation, though its predictive ability underscores the need for more robust biomarkers of treatment response.

## Data Availability

All data produced in the present study are available upon reasonable request to the authors

## Figure legends

**Supplementary Figure 1**. A flow diagram demonstrating the patient inclusion and exclusion criteria as well as the sample splitting procedure.

**Supplementary Figure 2**. A graph depicting the Hosmer-Lemeshow model calibration test, which demonstrates good logistic regression model calibration.

