## Supplementary Figure 1 for "Development of a clinical model to predict vagus nerve stimulation response in pediatric patients with drug-resistant epilepsy"

Retrospective collection of all patients with ASM-resistant epilepsy who underwent VNS surgery at CHP between Jan. 2009 and Dec. 2020 (n=423)

Patients excluded if age at VNS implantation >21 years

Validation sample (n=110)

*n=103 with outcome data*

Training sample (n=255)

*n=236 with outcome data*

Final total sample (n=365)

*n=339 with outcome data*

Random 70%

Random 30%
