## Supplementary figures and images for "Development of a clinical model to predict vagus nerve stimulation response in pediatric patients with drug-resistant epilepsy"

### Supplementary Figure 2

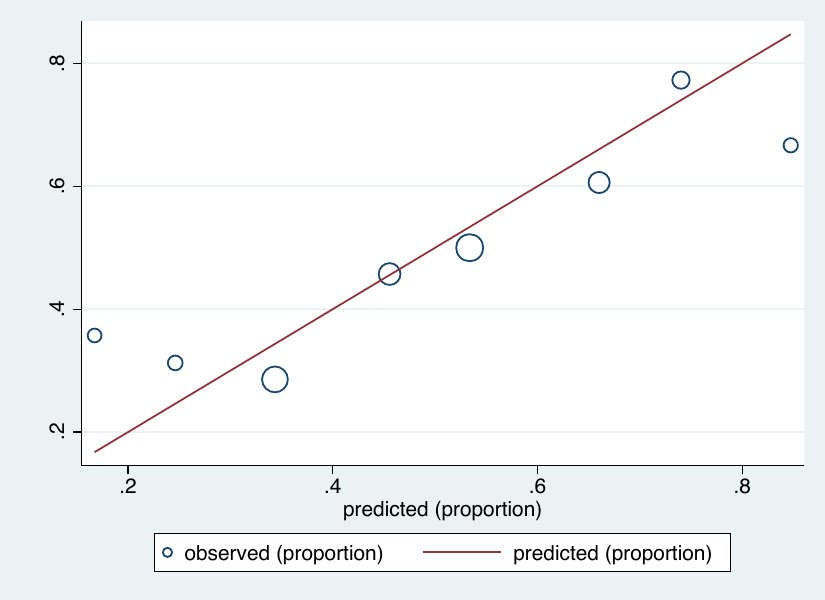
