## Supplementary Table 1 for "Development of a clinical model to predict vagus nerve stimulation response in pediatric patients with drug-resistant epilepsy"

|  | OR (95% confidence interval) | p-value |
| --- | --- | --- |
| Duration of epilepsy pre-VNS | | |
| >12 years* | 1 | -- |
| 9-12 years | 1.48 (0.56-3.92) | 0.430 |
| 4-8 years | 1.31 (0.58-2.99) | 0.516 |
| 0-3 years | 2.90 (1.19-7.08) | **0.019** |
| Age at seizure onset | | |
| <3 years* | 1 | -- |
| ≥3 years | 1.63 (1.06-2.52) | **0.028** |
| # of pre-VNS ASMs | | |
| 4-5* | 1 | -- |
| 2-3 | 2.19 (0.82-5.88) | 0.119 |
| 0-1 | 2.78 (0.94-8.27) | 0.066 |
| Semiology | | |
| Generalized non-motor* | 1 | **--** |
| Generalized motor | 1.34 (0.61-2.95) | 0.464 |
| Focal to tonic clonic | 2.16 (0.86-5.44) | 0.101 |
| Focal non-motor | 1.63 (0.68-3.92) | 0.277 |
| Focal motor | 1.87 (0.83-4.20) | 0.132 |
| Etiology^+^ | | |
| Unknown* | 1 | **--** |
| Structural | 1.04 (0.62-1.74) | 0.880 |
| Structural | 1.02 (0.59-1.75) | 0.954 |
| Infectious | 4.05 (0.81-20.29) | 0.089 |
| First therapeutic epilepsy surgery | | |
| Not VNS* | 1 | -- |
| VNS | 1.29 (0.51-3.30) | 0.592 |
| Supplemental Table 1. Odds ratios (ORs) and p-values from univariate logistic regressions evaluating the association between respective covariates and VNS response at one year, fitted on the total sample. Higher ORs are indicative of higher odds of VNS response. VNS: vagus nerve stimulation, ASMs: antiseizure medications. p<0.05 was considered significant.  *designates reference category  ^+^of the 3 patients in our sample with metabolic epilepsy, none had 1-year outcome data. Metabolic is not, therefore, an etiology presented in this table. | | |
