## Supplementary Table 2 for "Development of a clinical model to predict vagus nerve stimulation response in pediatric patients with drug-resistant epilepsy"

| Point total | n | Predicted response | Actual response |
| --- | --- | --- | --- |
| 0 | 0 | 0.03 | -- |
| 1 | 9 | 0.09 | 0.44 |
| 2 | 67 | 0.22 | 0.40 |
| 3 | 136 | 0.46 | 0.43 |
| 4 | 84 | 0.72 | 0.55 |
| 5 | 14 | 0.88 | 0.93 |
| Supplementary Table 2. Within our total sample for each point total, the overall sample size (n), the score’s predicted probability of VNS response at one year, and the observed proportion of patients who responded to VNS at one year. | | | |
